# First and second SARS-CoV-2 waves in inner London: A comparison of admission characteristics and the impact of the B.1.1.7 variant

**DOI:** 10.1101/2021.03.16.21253377

**Authors:** L.B. Snell, W. Wang, A. Alcolea-Medina, T. Charalampous, G. Nebbia, R. Batra, L. de Jongh, F. Higgins, Y. Wang, J.D. Edgeworth, V. Curcin

## Abstract

**Introduction:** A second wave of SARS-CoV-2 infection spread across the UK in 2020 linked with emergence of the more transmissible B.1.1.7 variant. The emergence of new variants, particularly during relaxation of social distancing policies and implementation of mass vaccination, highlights the need for real-time integration of detailed patient clinical data alongside pathogen genomic data. We linked clinical data with viral genome sequence data to compare cases admitted during the first and second waves of SARS-CoV-2 infection.

**Methods:** Clinical, laboratory and demographic data from five electronic health record (EHR) systems was collected for all cases with a positive SARS-CoV-2 RNA test between March 13th 2020 and February 17th 2021. SARS-CoV-2 viral sequencing was performed using Oxford Nanopore Technology. Descriptive data are presented comparing cases between waves, and between cases of B.1.1.7 and non-B.1.1.7 variants.

**Results:** There were 5810 SARS-CoV-2 RNA positive cases comprising inpatients (n=2341), healthcare workers (n=1549), outpatients (n=874), emergency department (ED) attenders not subsequently admitted (n=532), inter-hospital transfers (n=281) and nosocomial cases (n=233). There were two dominant waves of hospital admissions, with wave one starting from March 13^th^ (n=838) and wave two from October 20^th^ (n=1503), both with a temporally aligned rise in nosocomial cases (n=96 in wave one, n=137 in wave two). 1470 SARS-CoV-2 isolates were successfully sequenced, including 216/838 (26%) admitted cases from wave one, 472/1503 (31%) admitted cases in wave two and 121/233 (52%) nosocomial cases. The first B.1.1.7 variant was identified on 15th November 2020 and increased rapidly such that it comprised 400/472 (85%) of sequenced isolates from admitted cases in wave two. Females made up a larger proportion of admitted cases in wave two (47.3% vs 41.8%, p=0.011), and in those infected with the B.1.1.7 variant compared to non-B.1.1.7 variants (48.0% vs 41.8%, p=0.042). A diagnosis of frailty was less common in wave two (11.5% v 22.8%, p<0.001) and in the group infected with B.1.1.7 (14.5% v 22.4%, p=0.001). There was no difference in severity on admission between waves, as measured by hypoxia at admission (wave one: 64.3% vs wave two: 65.5%, p=0.67). However, a higher proportion of cases infected with the B.1.1.7 variant were hypoxic on admission compared to other variants (70.0% vs 62.5%, p=0.029).

**Conclusions:** Automated EHR data extraction linked with SARS-CoV-2 genome sequence data provides valuable insight into the evolving characteristics of cases admitted to hospital with COVID-19. The proportion of cases with hypoxia on admission was greater in those infected with the B.1.1.7 variant, which supports evidence the B.1.1.7 variant is associated with more severe disease. The number of nosocomial cases was similar in both waves despite introduction of many infection control interventions before wave two, an observation requiring further investigation.

## Background

SARS-CoV-2 infection has led to the death of over 1 million individuals worldwide since its emergence in China during December 2019, with over 100,000 deaths reported in the UK. SARS-CoV-2 incidence shows a highly dynamic course often described in waves. The first wave involved community and nosocomial transmission following the introduction of SARS-CoV-2 into a non-immune population. In London, the estimated incidence in the first wave peaked around March 23rd at 2.2% [1] and then rapidly declined following 2 weeks of social distancing and restrictions. Hospital admissions peaked about 1 week later [2] reflecting the median period of symptoms before hospital presentation.

Incidence of SARS-CoV-2 remained low during the summer months in the UK before a “second wave” of infections, starting in London around the beginning of October [3]. The second wave occurred in a very different environment. Many public health interventions had been introduced such as universal mask use in hospitals [4] and many indoor settings [5]. There was better understanding of risk factors for severe disease [6,7], which prompted vulnerable and elderly people to shield. People were urged to work from home when possible [8]. Schools reopened in September and students returned to University probably representing the largest population movement around the country preceding the second wave [9]. There was also rapid scale up of community testing with contact tracing and isolation. Genome sequencing identified new variants, including the B.1.1.7 variant which was first identified around the South East of England and spread rapidly as part of the emerging second wave. [10] The B.1.1.7 variant has been associated with increased transmissibility in community studies [11] [12] [13], and there is evidence from community studies that it may also confer increased mortality [14] [15] [16] [17]. Finally, vaccination is being rolled out at pace prioritising high risk groups and healthcare workers (HCW), with over 25% of the adult population receiving the first dose by mid February 2021 [18]

These many changes have potential to affect the burden on healthcare systems, and modify the characteristics of cases presenting to hospital including the demographics, co-morbidities and severity of disease associated with SARS-CoV-2 infection. For instance, a report from Japan suggests cases admitted during the second wave are younger, with fewer co-morbidities and with lower markers of disease severity [19]. It is important to understand these changes in real time, to help assess the impact of public health interventions and predict the potential burden on the healthcare system to plan resourcing and potentially modify community interventions.

Currently data are collected in routine clinical systems, alongside which ISARIC WHO CCP-UK collects data as a large prospective cohort study of COVID-19 patients [20] and COG-UK [21] supports collection of isolates for genome sequencing. As SARS-CoV-2 prevalence falls to potentially become endemic, informatics teams would benefit from an automated mechanism to iteratively extract and link patient and SARS-CoV-2 genomic data for local healthcare planning and to support national and international research studies. We therefore established a dataset integrating multiple health record systems and linked with local SARS-CoV-2 variant analysis to compare admission characteristics of hospitalised cases during the two dominant waves of infection.

## Methods

### Population of interest and setting

Guy’s and St Thomas’ NHS Foundation Trust (GSTT) is a multi-site inner-city healthcare institution providing general and emergency services predominantly to the South London boroughs of Lambeth and Southwark. The acute-admitting site (St Thomas’ Hospital) has an adult emergency department, with a large critical care service including one of the UK’s eight nationally commissioned ECMO centres for severe respiratory failure. A second site (Guy’s Hospital) provides elective surgery, cancer care and other specialist services. A paediatric hospital (Evelina London) and several satellite sites for specialist services like dialysis, rehabilitation and long term care are also part of the Trust. COVID-19 cases are admitted through different pathways comprising emergency department (ED), tertiary referrals to the severe respiratory failure service (ECMO), through specialist referral pathways and clinics (eg haemato-oncology and renal transplantation), and from regional hospitals predominantly to critical care through ‘mutual aid’ scheme.

### SARS-CoV-2 laboratory testing

GSTT has an on-site laboratory providing SARS-CoV-2 testing to all patients and HCW. The policies and technologies employed for SARS-CoV-2 testing changed over time based on national and local screening guidance and improvements in diagnostics. Our laboratory began testing on 13^th^ March 2020 with initial capacity for around 150 tests per day, before increasing to around 500 tests per day in late April during wave one, and up to 1000 tests per day during wave two (Supplementary Figure 1)

Assays used for the detection of SARS-CoV-2 RNA include PCR testing using Aus Diagnostics or by the Hologic Aptima SARS-CoV-2 Assay. Nucleic acid was first extracted using the QIAGEN QIAsymphony SP system and a QIAsymphony DSP Virus/Pathogen Mini Kit (catalogue No: 937036) with the off-board lysis protocol. This method utilises 200µl respiratory specimen with a 60µl elution volume. Testing commenced during the first wave on 13th March 2020 limited to cases requiring admission or inpatients who had symptoms of fever or cough, as per national recommendation; guidance suggested cases who did not require admission should not be tested. For wave two, all cases admitted to hospital were screened and underwent universal interval screening at varying time points. Staff testing for symptomatic healthcare workers was also introduced towards the end of wave one. Comparative analysis was therefore restricted to SARS-CoV-2 RNA positive cases requiring admission. Cases without laboratory confirmation of SARS-CoV-2 infection were not included.

### Definitions

Cases were identified by first positive SARS-CoV-2 RNA test. Cases were placed in mutually exclusive categories with the following definitions: 1) outpatients 2) testing through occupational health 3) emergency department attenders not subsequently admitted within 14 days 4) patients admitted within 14 days of a positive test 5) nosocomial cases, defined based on ECDC definitions, as those having a first positive test on day 8 or later after admission to hospital where COVID-19 was not suspected on admission [22] and 6) interhospital transfers. For the purpose of comparison only the inpatient group, admitted within 14 days following a positive test, were taken forward for onward comparison. A composite datapoint for ‘hypoxia’ was created, with cases taken to be hypoxic if on admission they had oxygen saturations of <94%, if they were recorded as requiring supplemental oxygen, or if the fraction of inspired oxygen was recorded as being greater than 0.21.

### Determination of SARS-CoV-2 lineage

Whole genome sequencing of residual samples from SARS-CoV-2 cases was performed using GridION (Oxford Nanopore Technology), using version 3 of the ARTIC protocol [23] and bioinformatics pipeline [24]. Samples were selected for sequencing if the corrected CT value was 33 or below, or the Hologic Aptima assay was above 1000 RLU. During the first wave sequencing occurred between March 1st - 31st, whilst sequencing in the second wave restarted in November 2020 and is ongoing. Lineage determination was performed using updated versions of pangolin 2.0 [25]. Samples were regarded as successfully sequenced if over 50% of the genome was recovered and if lineage assignment by pangolin was given with at least 50% confidence.

### Data sources, extraction and integration

Clinical, laboratory and demographic data for all cases with a laboratory reported SARS-CoV-2 PCR RNA test on nose and throat swabs or lower respiratory tract specimens were extracted from hospital electronic health record (EHR) data sources using records closest to the test date (DXC Technology’s i.CM EPR, Philips IntelliVue Clinical Information Portfolio (ICIP) Critical Care, DXC Technology’s MedChart, e-Noting and Citrix Remote PACS - Sectra). Data was linked to the Index of Multiple Deprivation (IMD), with 1 denoting the least deprived areas, and 5 the most deprived ones. Age and sex were extracted from EPR. Self-reported ethnicity of cases were stratified according to the 18 ONS categories of White (British, Irish, Gypsy and White-Other), Black (African, Caribbean, and Black-Other), Asian (Bangladeshi, Chinese, Indian, Pakistan, and Asian-Other), and Mixed/Other, with cases from Black ethnicities further separated into Black-Caribbean and Black-African.

Comorbidities and medication history were extracted from the EPR and e-Noting using natural language processing (NLP). Comorbidities were extracted from any of the databases covering the pathway of the cases from arrival in accident and emergency through inpatient general ward and critical care unit, where applicable, to hospital discharge or death. If a comorbidity was not recorded we assume that it was not present. Cases were characterised as having/not having a past medical history of hypertension, cardiovascular disease (stroke, transient ischaemic attack, atrial fibrillation, congestive heart failure, ischaemic heart disease, peripheral artery disease or atherosclerotic disease), diabetes mellitus, chronic kidney disease, chronic respiratory disease (chronic obstructive pulmonary disease, asthma, bronchiectasis or pulmonary fibrosis) and neoplastic disease (solid tumours, haematological neoplasias or metastatic disease). Obesity was defined as either morbid obesity present in the notes, or recorded Body Mass Index (BMI) >= 30 kg/m^2^.

Data management was performed using SQL databases, with analysis carried out on the secure King’s Health Partners (KHP) Rosalind high-performance computer infrastructure [26] running Jupyter Notebook 6.0.3, R 3.6.3 and Python 3.7.6. Medicines data were extracted using both structured queries and natural language processing tools with medical and drug dictionaries. Additionally, checks on free text data were performed by a cardiovascular clinician to ensure the information was accurate.

### Statistical analysis

The statistical analysis in the paper is mainly descriptive. The general statistics were summarised with mean and standard deviation (SD) for continuous variables if the distribution is normal and median and interquartile range (IQR) if the distribution is non-normal. Count and percentages were used for categorical variables. For the comparisons between wave one and two variables, Kruskal-Walllis test was used for continuous variables and Chi-squared test for categorical variables. The reference significant level was set to be p=0.05.

## Results

### General epidemiology and results of viral sequencing

Figure 1 shows the incidence of SARS-CoV-2 cases, SARS-CoV-2 admissions, and nosocomial cases since March 13th 2020. In total 5810 individuals had a positive SARS-CoV-2 PCR test up until the data extraction date of 17th February 2021. Two “waves” are evident with July 25th taken as an arbitrary separation date between waves, at which point a minimum of 12 wave one cases remained in hospital. Wave one comprised 1528 unique cases (26.3%) from when laboratory testing commenced on March 13th to peak rapidly between the 1st and 8th April 2020 with 57 new cases, before falling to a baseline by May 12th 2020. 1391/1528 (91%) of all cases in wave one occurred during these 60 days. Wave two comprised 4282 unique cases (73.7%), with incidence first increasing gradually from the beginning of October. There was then a period of rapidly escalating incidence from about 10th December, peaking on 28th December 2020 139 cases were diagnosed. 3446/4282 (80%) of wave two cases detected during a comparable 60 day period ending 8th February 2021. The number of cases admitted, the primary focus of this report, and the number of nosocomial cases are highlighted in Figure 1. In both waves nosocomial cases peaked early increasing along with admissions but then fell while the number of community admissions continued at peak levels.

**Figure 1:**
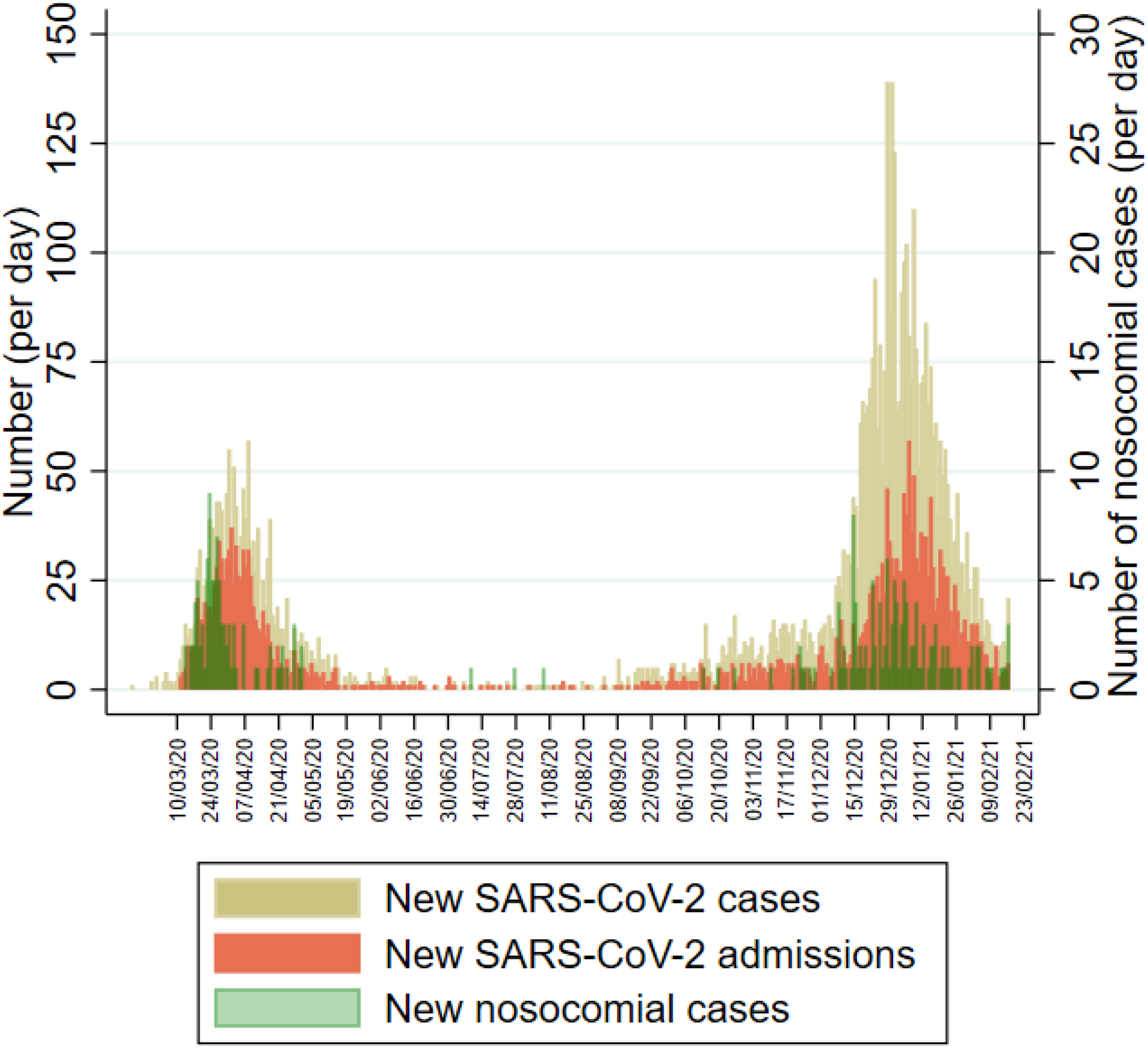
Distribution of laboratory confirmed SARS-CoV-2 cases over time. Daily incidence of new cases (beige), newly admitted cases (orange) and nosocomial acquisitions (green) over time.

Individuals with a positive test were placed into six categories (Figure 2). Some observed differences between wave one and two reflected the increased availability of testing particularly for outpatients (208;13.6% v 666;15.6%), people sent home from ED (111;7.3% v 421; 9.8%) and healthcare workers (171;11.2% v 1378;32.2%). There were also more interhospital transfers of known COVID-19 cases in wave two (177;4.1% v 104;6.8% in wave one). In wave two, the number of admissions increased (1503; 35.1% v 838; 54.8%) along with nosocomial cases (137;3.2% v 96;6.3%) compared with wave one.

**Figure 2.**
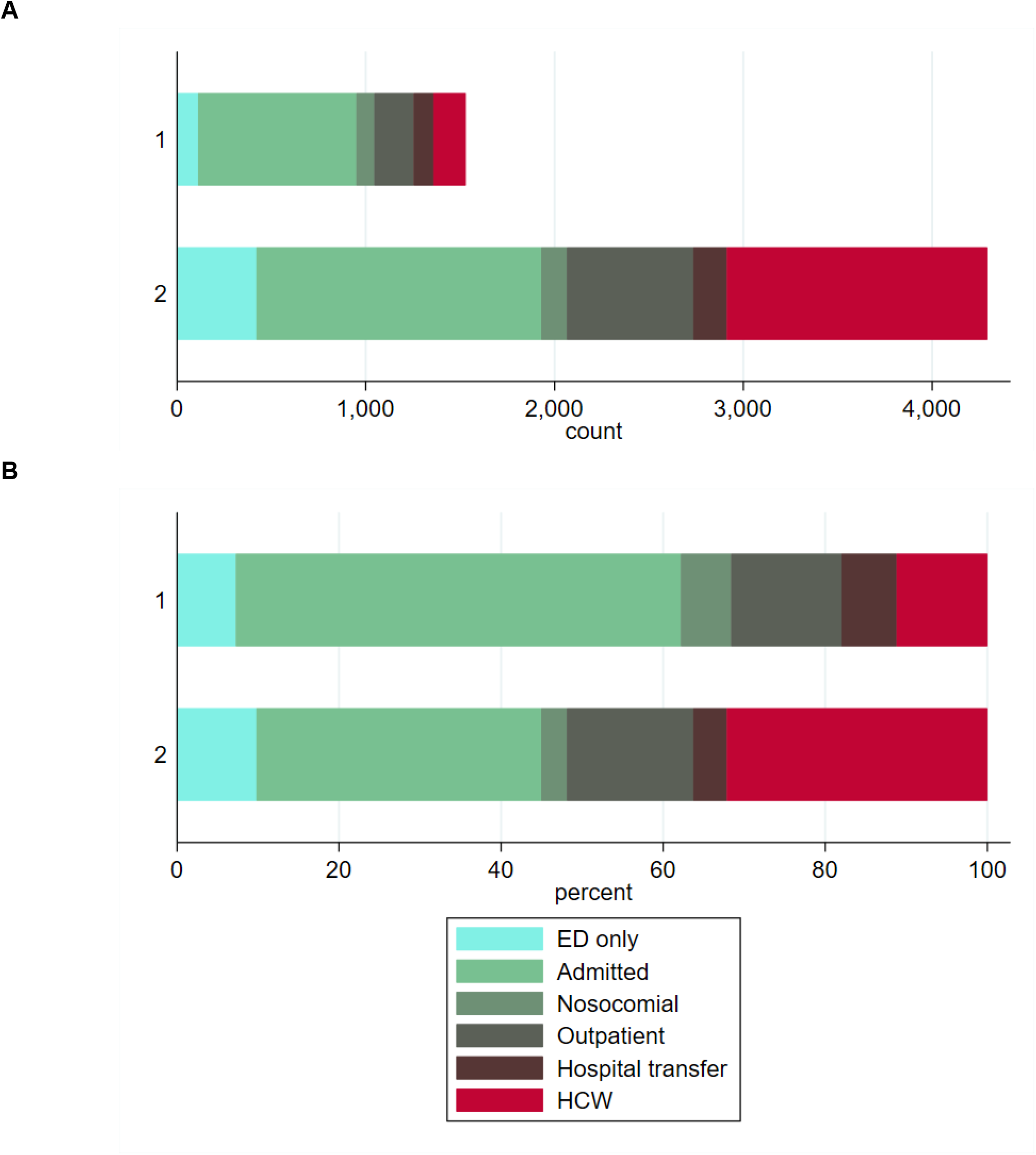
**(A)** Absolute number of cases within the different hospital cohorts during wave one (upper) and wave two (lower). **(B)** Proportion of cases within the different hospital cohorts during wave one (upper) and wave two (lower).

Figure 3 shows the number of successfully sequenced SARS-CoV-2 isolates over time, with 382 from wave one and 1088 from wave two. The proportion of B.1.1.7 variant increased rapidly after the first B.1.1.7 isolate was identified on 15th November 2020, accounting for approximately two thirds within 3 weeks, and almost 100% (600/617 isolates, 97%) in January 2021. In the second wave, the B.1.1.7 variant made up 83% (908/1088) of all sequenced isolates, 85% (400/472) of sequenced isolates from admitted cases, and 88% (51/59) of sequenced isolates from nosocomial cases. In addition, two cases of the B.1.351 variant of concern were also detected in the wave two admission cohort.

**Figure 3.**
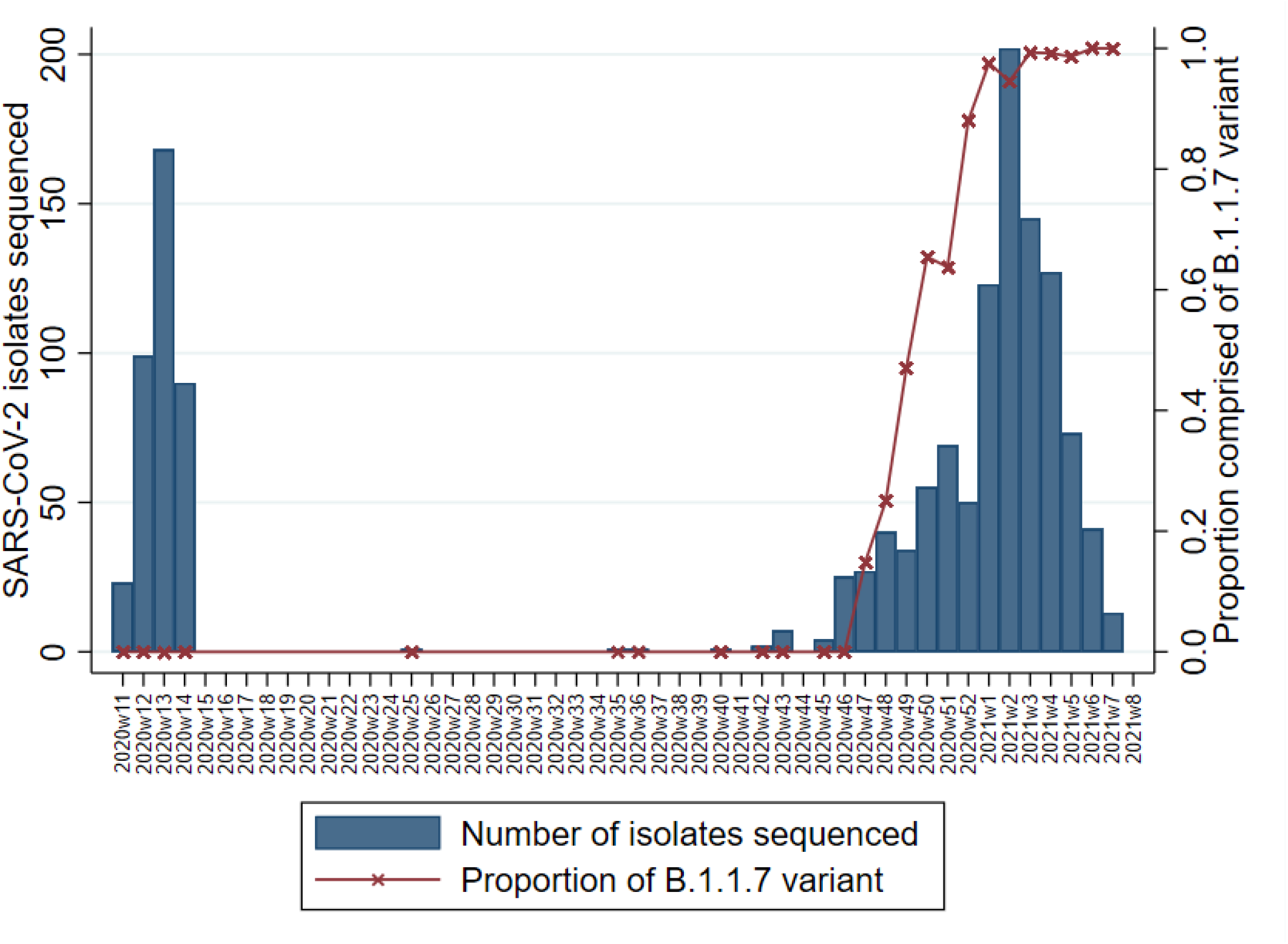
Number of cases with sequenced SARS-CoV-2 isolates by epi-week (bar) and the proportion of which were made up of the variant B.1.1.7 (red line)

### Comparison of characteristics of admitted cases between wave one and two

General statistics of cases admitted during wave one (n=838) and wave two (n=1503) were compared (Table 1). There was only a small difference in mean age (62yrs in wave one v 60yrs in wave two, p=0.019), however admitted cases were more likely to be female in wave two (47.3% v 41.8%, p=0.011). Comparison of comorbidities showed those in wave two were less likely to have a diagnosis of frailty (11.5% v 22.8%, p<0.001), history of stroke (4.3% v 8.6%, p<0.001) or cancer (4.8% v 7.2%, p=0.022). A larger proportion of admitted cases in wave two were obese (29.1% v 24.6%, p=0.02). There was no significant difference in proportion with known comorbidities of diabetes, kidney disease, hypertension, cardiovascular disease or respiratory disease.

**Table 1.** General statistics of the cohort for wave one and two admissions.

|  | Missing | Wave one admissions<br>n (%) / value [IQR] | Wave two admissions<br>n (%) / value [IQR] | P-Value |
| --- | --- | --- | --- | --- |
| n |  | 838 | 1503 |  |
| Demographics |  |  |  |  |
| Age | 0 | 62.0 [49.0,78.0] | 60.0 [47.0,74.0] | 0.019 |
| Male | 0 | 488 (58.2) | 792 (52.7) | 0.011 |
| Ethnicity | 0 |  |  | 0.013 |
| White |  | 331 (39.5) | 598 (39.8) |  |
| Asian |  | 64 (7.6) | 121 (8.1) |  |
| Black-African |  | 177 (21.1) | 262 (17.4) |  |
| Black-Caribbean |  | 73 (8.7) | 98 (6.5) |  |
| Mixed |  | 15 (1.8) | 18 (1.2) |  |
| Other |  | 45 (5.4) | 107 (7.1) |  |
| Unknown |  | 133 (15.9) | 299 (19.9) |  |
| BMI | 577 | 27.0 [23.8,31.7] | 27.7 [24.0,32.9] | 0.022 |
| BMI>30 |  | 206 (24.6) | 438 (29.1) | 0.02 |
| BMI>40 |  | 34 (4.1) | 86 (5.7) | 0.098 |
| Physiological parameters |  |  |  |  |
| Heart Rate | 360 | 84.0 [75.0,94.0] | 81.0 [72.0,91.0] | <0.001 |
| Heart Rate>100 |  | 105 (12.5) | 142 (9.4) | 0.02 |
| Blood pressure |  |  |  |  |
| Systolic | 369 | 125.0 [113.0,139.0] | 127.0 [115.0,141.0] | 0.013 |
| Diastolic | 369 | 73.0 [65.0,80.0] | 75.0 [68.0,82.0] | <0.001 |
| MAP | 369 | 90.7 [82.2,99.0] | 92.3 [84.7,101.3] | <0.001 |
| Respiratory Rate | 359 | 19.0 [18.0,22.0] | 19.0 [18.0,22.0] | 0.764 |
| Respiratory Rate>20 |  | 200 (23.9) | 365 (24.3) | 0.86 |
| Hypoxia | 658 | 370 (64.3) | 726 (65.5) | 0.67 |
| Temperature | 361 | 36.9 [36.4,37.5] | 36.6 [36.2,37.2] | <0.001 |
| NEWS2 | 405 |  |  | 0.86 |
| 0 |  | 95 (11.3) | 173 (11.5) |  |
| 1 |  | 108 (12.9) | 192 (12.8) |  |
| 2 |  | 117 (14.0) | 188 (12.5) |  |

| 2+ |  | 371 (44.3) | 692 (46.0) |  |
| --- | --- | --- | --- | --- |
| Laboratory parameters |  |  |  |  |
| Neutrophils | 8 | 4.9 [3.4,7.6] | 5.0 [3.3,7.5] | 0.724 |
| Lymphocytes | 7 | 0.9 [0.6,1.3] | 0.9 [0.6,1.4] | 0.741 |
| NLR | 8 | 5.4 [3.1,9.9] | 5.4 [3.2,9.8] | 0.951 |
| Creatinine | 43 | 83.0 [64.0,115.0] | 86.0 [68.0,117.0] | 0.065 |
| Urea | 855 | 7.0 [4.6,12.2] | 6.0 [4.3,9.9] | 0.001 |
| Estimated GFR | 114 | 73.0 [48.0,98.0] | 69.0 [48.0,89.0] | 0.001 |
| Albumin | 185 | 37.0 [32.0,40.0] | 38.0 [34.0,41.0] | <0.001 |
| CRP | 61 | 74.5 [26.0,148.0] | 51.0 [18.0,103.8] | <0.001 |
| DDimer | 1297 | 1.1 [0.6,3.0] | 0.9 [0.5,2.2] | 0.001 |
| Ferritin | 905 | 855.0 [394.0,1533.5] | 699.0 [342.0,1359.0] | 0.05 |
| Co-morbidities |  |  |  |  |
| Stroke | 0 | 72 (8.6) | 64 (4.3) | <0.001 |
| TIA | 0 | 9 (1.1) | 20 (1.3) | 0.731 |
| Hypertension | 0 | 288 (34.4) | 464 (30.9) | 0.091 |
| Diabetes | 0 | 246 (29.4) | 384 (25.5) | 0.052 |
| AF | 0 | 63 (7.5) | 115 (7.7) | 0.972 |
| IHD | 0 | 146 (17.4) | 244 (16.2) | 0.495 |
| Heart Failure | 0 | 54 (6.4) | 105 (7.0) | 0.679 |
| COPD | 0 | 64 (7.6) | 109 (7.3) | 0.796 |
| Asthma | 0 | 74 (8.8) | 138 (9.2) | 0.835 |
| Cancer | 0 | 60 (7.2) | 72 (4.8) | 0.022 |
| Kidney disease | 0 | 112 (13.4) | 181 (12.0) | 0.389 |
| HIV | 0 | 21 (2.5) | 36 (2.4) | 0.979 |
| Solid organ Transplant | 0 | 24 (2.9) | 49 (3.3) | 0.686 |
| Frailty | 0 | 191 (22.8) | 173 (11.5) | <0.001 |

Note: p-value was from Kruskal-Wallis test for continuous variables and Chi-squared test for categorical variables. Abbreviations: BMI: Body Mass Index, TIA: Transient Ischaemic Attack, IHD: Ischemic Heart Disease, COPD: Chronic Obstructive Pulmonary Disease, HIV: Human Immunodeficiency Virus, MAP: Mean Arterial Pressure, SPO2: Oxygen Saturation, GFR: Glomerular Filtration Rate, CRP: C-Reactive Protein, NLR: Neutrophils and Lymphocytes Ratio, NEWS2: National Early Warning Score 2.

There were no significant differences between waves in the proportion with severe SARS-CoV-2 disease upon admission as judged by hypoxia (64.3% in wave one vs 65.5% in wave two, p=0.67) or tachypnoea (respiratory rate>20) (23.9% vs 24.3%, p=0.86). There were small differences in other physiological parameters on admission, some of which reached statistical significance but differences were not clinically relevant.

Laboratory markers were compared between waves (Table 1). There were small but significant differences, such as lower CRP (median 51.0 mg/dL (IQR: 18.0-103.8) v 74.5 mg/dL (IQR: 26.0-148.0), p<0.001) and lower ferritin (699.0 [IQR: 342.0-1359.0] v 855.0 [IQR: 394.0-1533.5], p=0.05) in wave two. There were other small statistically significant differences without clear clinical significance, such as a lower D-Dimer in wave two (0.9 mg/L FEU [IQR: 0.5-2.2] v 1.1 mg/L FEU [IQR:0.6-3.0], p=0.001) and lower estimated GFR (69.0 ml/min [IQR: 48.0-89.0] v 73.0 ml/min [IQR: 48.0-98.0], p=0.001), lower urea (6.0 mmol/L [IQR: 4.3-9.3] v 7.0 mmol/L [IQR: 4.6-12.2], p=0.001) and higher albumin (38.0 g/L [IQR: 34.0-41.0 g/L] vs 37.0 g/L [IQR: 32.0-40.0], p<0.001). There was no significant difference with neutrophils, lymphocytes, neutrophils and lymphocytes ratio (NLR), creatinine and glucose.

### Comparison of characteristics of admitted cases infected with B.1.1.7 and non-B.1.17 variants

Given the reported association between increased disease severity and transmission with the B.1.1.7 variant, we compared demographic, physiological and laboratory parameters between admitted cases with infection caused by B.1.1.7 variant (n=400) compared with non-B.1.1.7 (n=910) variants (Table 2). We considered all cases in wave one to be non-B.1.1.7, as wave one took place prior to emergence of the B.1.1.7 variant and before B.1.1.7 variant was first identified in our population in November 2021. Groups with non-B.1.1.7 and B.1.1.7 variant were not significantly different in mean age (62yrs vs 64yrs, p=0.22) or ethnicity. The proportion of admissions who were female was larger in the group infected with the B.1.1.7 variant compared to those infected by non-B.1.1.7 variants (48.0% vs 41.8%, p=0.01).

**Table 2.** General statistics of the cohort for non B.1.1.7 variant and B.1.1.7 variant admissions.

|  | Missing | Non B.1.1.7 variant<br>n (%) / value [IQR] | B.1.1.7 variant<br>n (%) / value [IQR] | P-Value |
| --- | --- | --- | --- | --- |
| n |  | 910 | 400 |  |
| Demographics |  |  |  |  |
| Age | 0 | 62.0 [49.0,78.0] | 64.0 [52.0,78.0] | 0.22 |
| Male |  | 530 (58.2) | 208 (52.0) | 0.042 |
| Ethnicity | 0 |  |  | 0.402 |
| White |  | 358 (39.3) | 164 (41.0) |  |
| Asian |  | 71 (7.8) | 38 (9.5) |  |
| Black-African |  | 191 (21.0) | 67 (16.8) |  |
| Black-Caribbean |  | 78 (8.6) | 27 (6.8) |  |
| Mixed |  | 16 (1.8) | 6 (1.5) |  |
| Other |  | 50 (5.5) | 23 (5.8) |  |
| Unknown |  | 146 (16.0) | 75 (18.8) |  |
| BMI | 334 | 27.1 [23.8,31.7] | 28.1 [24.0,34.2] | 0.036 |
| BMI>30 |  | 226 (24.8) | 121 (30.2) | 0.048 |
| BMI>40 |  | 36 (4.0) | 26 (6.5) | 0.063 |
| Physiological parameters |  |  |  |  |
| Heart Rate | 198 | 84.0 [74.0,94.0] | 80.0 [72.0,90.0] | 0.001 |
| Heart Rate>100 |  | 118 (13.0) | 36 (9.0) | 0.05 |
| Blood pressure |  |  |  |  |
| Systolic | 201 | 125.0 [113.0,139.5] | 127.0 [115.0,142.0] | 0.138 |
| Diastolic | 201 | 73.0 [65.0,80.0] | 75.0 [67.0,83.0] | 0.01 |
| MAP | 201 | 90.7 [82.3,99.2] | 92.7 [84.0,101.7] | 0.022 |
| Respiratory Rate | 194 | 19.0 [18.0,21.0] | 19.0 [18.0,22.0] | 0.591 |
| Respiratory Rate>20 |  | 209 (23.0) | 96 (24.0) | 0.737 |
| Hypoxia | 0 | 392 (62.5) | 217 (70.0) | 0.029 |
| Temperature | 199 | 36.9 [36.4,37.5] | 36.6 [36.2,37.1] | <0.001 |
| NEWS2 | 0 |  |  | 0.038 |
| 0 |  | 107 (11.8) | 43 (10.8) |  |
| 1 |  | 125 (13.7) | 39 (9.8) |  |
| 2 |  | 127 (14.0) | 53 (13.2) |  |
| 2+ |  | 391 (43.0) | 207 (51.7) |  |
| nan |  | 160 (17.6) | 58 (14.5) |  |
| Laboratory parameters |  |  |  |  |
| Neutrophils | 2 | 4.9 [3.4,7.6] | 4.8 [3.3,6.9] | 0.479 |
| Lymphocytes | 1 | 0.9 [0.6,1.3] | 0.8 [0.5,1.2] | 0.005 |
| NLR | 2 | 5.4 [3.1,9.9] | 5.8 [3.5,10.2] | 0.195 |
| Creatinine | 16 | 83.0 [64.0,115.0] | 92.0 [74.0,126.0] | <0.001 |
| Urea | 536 | 6.8 [4.3,12.0] | 6.6 [4.4,10.6] | 0.573 |
| Estimated GFR | 43 | 73.0 [48.5,97.0] | 63.5 [44.0,81.0] | <0.001 |
| Albumin | 107 | 37.0 [33.0,41.0] | 38.0 [34.0,41.0] | 0.009 |
| CRP | 21 | 70.0 [25.0,142.0] | 54.0 [24.0,102.0] | <0.001 |
| DDimer | 727 | 1.1 [0.6,2.8] | 0.9 [0.5,1.9] | 0.019 |
| Ferritin | 501 | 815.0 [366.2,1499.0] | 712.0 [357.5,1294.0] | 0.341 |
| Co-morbidities |  |  |  |  |
| Stroke | 0 | 74 (8.1) | 22 (5.5) | 0.117 |
| TIA | 0 | 12 (1.3) | 5 (1.2) | 0.87 |
| Hypertension | 0 | 315 (34.6) | 144 (36.0) | 0.674 |
| Diabetes | 0 | 267 (29.3) | 106 (26.5) | 0.326 |
| AF | 0 | 72 (7.9) | 42 (10.5) | 0.154 |
| IHD | 0 | 162 (17.8) | 78 (19.5) | 0.513 |
| Heart Failure | 0 | 61 (6.7) | 34 (8.5) | 0.299 |
| COPD | 0 | 74 (8.1) | 32 (8.0) | 0.977 |
| Asthma | 0 | 84 (9.2) | 39 (9.8) | 0.846 |
| Cancer | 0 | 64 (7.0) | 21 (5.2) | 0.278 |
| Kidney disease | 0 | 122 (13.4) | 62 (15.5) | 0.359 |
| HIV | 0 | 22 (2.4) | 10 (2.5) | 0.916 |
| Solid organ transplant | 0 | 25 (2.7) | 19 (4.8) | 0.092 |
| Frailty | 0 | 204 (22.4) | 58 (14.5) | 0.001 |

Note: p-value was from Kruskal-Wallis test for continuous variables and Chi-squared test for categorical variables. Abbreviations: BMI: Body Mass Index, TIA: Transient Ischaemic Attack, IHD: Ischemic Heart Disease, COPD: Chronic Obstructive Pulmonary Disease, HIV: Human Immunodeficiency Virus, MAP: Mean Arterial Pressure, SPO2: Oxygen Saturation, GFR: Glomerular Filtration Rate, CRP: C-Reactive Protein, NLR: Neutrophils and Lymphocytes Ratio, NEWS2: National Early Warning Score 2.

Cases infected with the B.1.1.7 variant were less likely to be frail (14.5% vs 22.4% p=0.001). A higher proportion of those in the B.1.1.7 group were obese (30.2% v 24.8%, p=0.048). Other minor differences in comorbidities between groups are shown in Table 2, but did not reach statistical significance.

On admission a higher proportion of those infected with B.1.1.7 were hypoxic (70.0% vs 62.5%, p=0.029), the main indicator of severe disease. CRP on admission was lower in the B.1.1.7 group (54 mg/L IQR: 24.0-102.0) compared to those infected with non-B.1.1.7 variants (70 mg/L, IQR: 25.0-142.0 p<0.001). Differences in other laboratory parameters did not meet either statistical or clinical significance.

## Discussion

Current hospital-based data collection methods predominantly rely on manual data extraction or linkage of multiple electronic records, which is resource intensive. Here we investigated the utility of automated data extraction from electronic health records as applied to investigation of changes during the second wave linked with emergence of a dominant new variant (B.1.1.7). Our data from a large healthcare institution in one of the worst affected regions internationally provides a large dataset for in-depth comparison; for instance we report a similar number of cases as reported from a national observational cohort study from Japan [19].

There were threefold more SARS-CoV-2 RNA positive cases reported by the hospital laboratory in wave two. The interpretation of this increase must recognise the many changes in testing guidelines and testing capacity between the two waves. Testing capacity increased substantially throughout 2020 both in hospital and in the community. Partly due to these capacity limits, during wave one it was not local policy to offer testing to outpatients and those not requiring admission, instead relying on clinical diagnosis. Healthcare workers were not offered occupational health testing until the end of wave one. We therefore restricted comparison to admitted and nosocomial cases.

There were almost twice as many admitted cases in wave two compared with wave one (1503 v 838). This is consistent with a higher local community incidence as reported by the ONS infection survey with 3.5% of individuals in London infected in January 2021 [27], compared with 2.2% of individuals in London at the peak of wave one [1]. The increase in peak hospital occupancy in wave two has also been reported nationally [28].

A major contributor to this increase in hospital admissions is likely to be the emergence of the B.1.1.7 variant, which is reported to be more transmissible and more virulent [14] [15] [16] [17]. However, the published studies thus far are largely based on mortality in community populations, and data on hospitalised cohorts is lacking. Our finding that those admitted with infection by the B.1.1.7 variant are more likely to be hypoxic on admission, a key marker of severe disease, is consistent with increased virulence of the B.1.1.7 variant as reported in these studies.

There was also an increase in the proportion of females in the admitted cohort of wave two, and those infected with B.1.1.7, accounting for an extra 5% of admissions with SARS-CoV-2 infection. Unpublished data referenced by NERVTAG [29] suggests hospitalised females infected with the B.1.1.7 variant may be more likely to die or require ICU care. Our data, showing an increase in the proportion of females in the admission cohort is consistent with the finding that B.1.1.7 may show increased virulence in females. Further analysis is required to determine whether this effect is attenuated when adjusted for age and other variables.

Admitted cases in the wave two were also around half as likely to have a diagnosis of frailty, which may be due to fewer admissions from care homes during wave two, which has been reported both nationally [30] and internationally [31]. Additionally, admitted cases were around a third less likely to have cancer in wave two. Both of these reductions may also be as a result of individuals shielding, and therefore at reduced risk for acquiring SARS-CoV-2 infection. Other differences in comorbidities between waves were small and of unclear clinical significance. Those admitted with B.1.1.7 infection were also less likely to be frail, however no other clinically significant difference in co-morbidities was found.

Our findings contrast to those seen in other areas where differences between waves have been compared. For instance, in Japan the second wave had fewer cases, which were younger and less comorbid [19]. Our results differ, showing a similar age distribution between waves, similar to what has been reported internationally [31] [32].

One additional striking observation was the similarity in number of nosocomial cases in wave one (n=96) and wave two (n=137). Interestingly, nosocomial cases in wave one increased and started to fall before impact of the main infection control interventions of banning hospital visitors (March 25th), introducing universal surgical mask wearing (28th March 2020) and universal regular inpatient screening (after the first wave). In comparison, all these measures were in place prior to the second wave. The increased number of cases in wave two may in part be due to increased inpatient screening, which would identify asymptomatic cases, or introduction of the more transmissible B.1.1.7 variant which made up the vast majority of our sequenced nosocomial cases.

In both waves the incidence of nosocomial cases rose temporally aligned with increasing community incidence and hospital admissions, and then started to fall while new admissions continued to increase. This is not consistent with the predominant mode by which nosocomial cases acquire infection being via transmission from admitted cases. It seems more likely that incidence in the community and in nosocomial cases are mechanistically linked and therefore follow the same course, peaking prior to hospitalisations of cases.

Some healthcare institutions report far fewer nosocomial acquisitions; for instance an academic hospital in Boston, USA reported only 2 nosocomial cases in over 9000 admissions [33]. This could be due to greater availability of side rooms for isolation or their use of N95 masks by HCWs, which may decrease transmission between HCWs and patients. In contrast, current UK public health policy recommends surgical facemasks for patient interactions unless performing aerosol generating procedures [34]. This incidence of nosocomial infection is a major challenge for UK healthcare institutions, with associated crude mortality at around 30% during the first wave [35,36]. For this reason it will be important to further investigate the factors involved in nosocomial acquisition in both waves.

### Limitations

Our study population comes from a single inner-city healthcare institution and therefore needs to be compared with findings from other centres. Our dataset included cases identified by SARS-CoV-2 RNA testing in our laboratory, so some cases diagnosed by external laboratories prior to admission may not be represented unless subsequently testing positive in our laboratory. The impact of differences in testing strategy and capacity during both waves needs further investigation, particularly the impact of the increase in asymptomatic cases in wave two. Finally, the analysis of clinical characteristics in this study was restricted to those recorded on admission to hospital. A follow-up study will include outcome data, when the wave two cohort has completed hospital stay, alongside other unstructured data points such as radiology or genome sequence results in clinical or external reports, to facilitate linkage with national and international research studies.

## Conclusion

Our data extraction method is able to iteratively extract clinical information from multiple clinical systems and integrate into a clinical dataset of SARS-CoV-2 cases. This compares to the more labour intensive prospective data collection by research staff conducting manual data collection. The study also highlights the benefits of being able to integrate detailed clinical data with genome sequence to obtain rapid insight into genotype/phenotype differences. The number of cases diagnosed, admissions and nosocomial cases were higher in wave two than wave one, likely due to the increased incidence caused by the more transmissible B.1.1.7 variant. A larger proportion of admitted cases in wave two and in those infected by B.1.1.7 were female. B.1.1.7 was associated with higher proportion being hypoxic on admission, which may reflect increased virulence of B.1.1.7.

## Data Availability

The original clinical and genomic datasets are not made available.

## Acknowledgements

The authors acknowledge use of the research computing facility at King’s College London, Rosalind (https://rosalind.kcl.ac.uk), which is delivered in partnership with the National Institute for Health Research (NIHR) Biomedical Research Centres at South London & Maudsley and Guy’s & St. Thomas’ NHS Foundation Trusts, and part-funded by capital equipment grants from the Maudsley Charity (award 980) and Guy’s & St. Thomas’ Charity (TR130505). The views expressed are those of the author(s) and not necessarily those of the NHS, the NIHR, King’s College London, or the Department of Health and Social Care.

## Figures and Tables

**Supplementary Figure 1:**
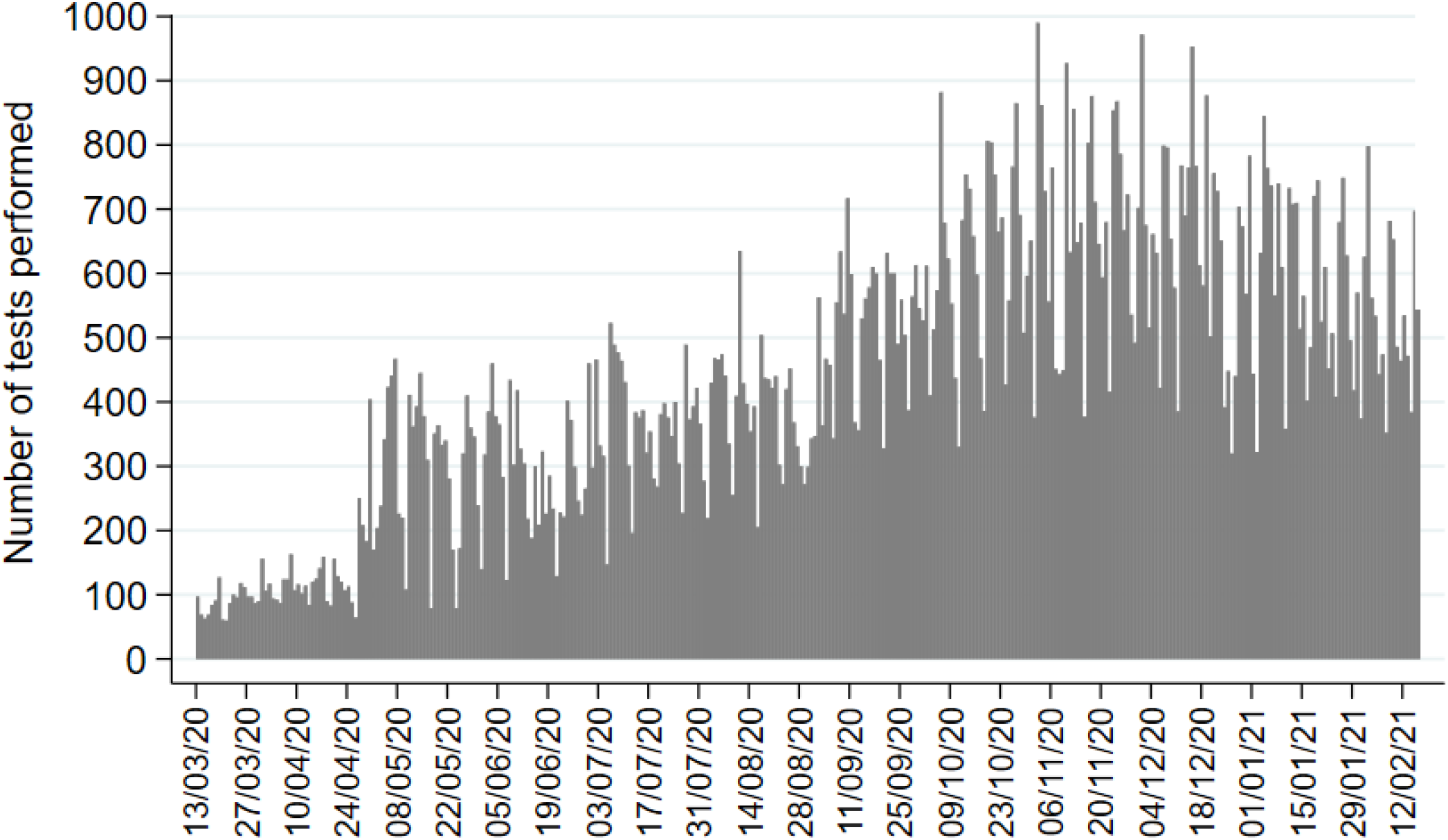
Daily number of SARS-CoV-2 tests performed in our laboratory between 13^th^ March 2021, when testing was introduced, until 17^th^ February 2021.

## Ethics

Ethical approval for data informatics was granted by The London Bromley Research Ethics Committee (reference (20/HRA/1871)) to the King’s Health Partners Data Analytics and Modelling COVID-19 Group to collect clinically relevant data points from patient’s electronic health records.

Whole genome sequencing of residual viral isolates was conducted under the COVID-19 Genomics UK (COG-UK) consortium study protocol, which was approved by the Public Health England Research Ethics and Governance Group (reference: R&D NR0195).

